# Genetic associations of adult height with risk of cardioembolic and other subtypes of ischaemic stroke: a Mendelian randomisation study in multiple ancestries

**DOI:** 10.1101/2020.08.26.20174086

**Authors:** Andrew B Linden, Robert Clarke, Jemma C Hopewell, Yu Guo, William N Whiteley, Kuang Lin, Iain Turnbull, Yiping Chen, Canqing Yu, Jun Lv, Alison Offer, Imen Hammami, Derrick Bennett, Robin G Walters, Liming Li, Zhengming Chen, Sarah Parish, for the China Kadoorie Biobank Collaborative Group

## Abstract

**Background:** Taller adult height is associated with lower risks of ischaemic heart disease in both observational and Mendelian randomisation studies, but little is known about the causal relevance of height for different subtypes of ischaemic stroke and the mechanisms involved.

**Methods:** Height-associated genetic variants (up to 2,931) from previous genome-wide association studies were used to construct genetic instruments in different populations. Two-sample Mendelian randomisation approaches were used to examine the associations of genetically-determined height with ischaemic stroke and its subtypes in multiple ancestries (MEGASTROKE: 60,341 ischaemic stroke cases) supported by additional cases in Europeans (UK Biobank: 4,055 cases) and in Chinese (China Kadoorie Biobank: 10,297 cases). The associations of genetically-determined height with established cardiovascular and other risk factors were also examined in Europeans (UK Biobank: 336,750 participants) and Chinese (China Kadoorie Biobank: 58,277 participants).

**Results:** Genetically-determined height was inversely associated with ischaemic stroke (4% [95% CI: 1–7] lower risk per 1 standard deviation taller height in MEGASTROKE). This masked much stronger opposing associations of height with different subtypes, with a 12% (95% CI: 6–17) higher risk of cardioembolic stroke, 11% (6–16) lower risk of large-artery stroke, and 14% (9–18) lower risk of small-vessel stroke. Genetically-determined height was strongly positively associated with atrial fibrillation, lean body mass and lung function, and inversely associated with levels of LDL cholesterol and blood pressure in both Europeans and Chinese.

**Conclusions:** In multiple ancestries, genetic associations support the causal relevance of taller adult height for higher risk of cardioembolic stroke (in addition to atrial fibrillation) and lower risk of other ischaemic strokes, highlighting the need to properly differentiate subtypes of ischaemic stroke in both clinical practice and research.

## Introduction

Taller people have lower risks of atherosclerotic diseases, ischaemic stroke and heart disease, but higher risks of atrial fibrillation and venous thromboembolism.^1,2^ The associations of height with ischaemic stroke subtypes have not been reported, but it would be of interest to know whether these vary between atherosclerotic and cardioembolic stroke subtypes. In observational studies, any such associations could reflect confounding by socioeconomic status or other known or unknown correlates of height that are risk factors for cardiovascular disease. Alternatively, the associations could be causal and could possibly be mediated through effects of height on body structure (including lean body mass or lung function).^3-6^

Increasingly Mendelian randomisation (MR) analyses have been used to assess the causal relevance of risk factors for diseases by using genetic variants associated with risk factors of interest as instrumental variables.^7^ The allocation of genetic variants to gametes (and hence offspring) is determined *randomly* at meiosis. Therefore, the random distribution of variants for a trait, such as height, between individuals can be used to minimise the effects of confounding by risk factors and provide support for the causal relevance of the trait for disease outcomes. Previous MR studies have reported that genetically-determined differences in adult height were inversely associated with ischaemic heart disease^3^ and hypertension, but positively associated with atrial fibrillation, venous thromboembolism, and vasculitis.^2^ However, the associations of genetically-determined height with ischaemic stroke and ischaemic stroke subtypes have not been reliably established as previous studies have been based primarily on total stroke rather than separately on stroke types and their main subtypes ^2,8^.

The present study examined the observational and genetic associations (using MR approaches) of height with: (i) ischaemic stroke and subtypes of ischaemic stroke in the MEGASTROKE consortium (an international collaboration on the genetics of stroke) and in two large prospective studies conducted in the UK and China,^9,10^ and (ii) established cardiovascular risk factors and anthropometric traits in the two large prospective studies.

## Methods

### MEGASTROKE

MEGASTROKE consortium data included 29 genome-wide studies of stroke and stroke subtypes.^11^ Ischaemic stroke cases were defined using standard diagnostic criteria based on clinical and imaging findings, and were further classified into subtypes using the Trial of ORG 10,172 in Acute Stroke Treatment (TOAST) criteria.^11,12^ Analyses were conducted using summary results from multiple ancestries (60,341 ischaemic stroke cases—including 9,006 cardioembolic stroke, 6,688 large-artery atherosclerotic stroke, and 11,710 small-vessel stroke subtypes—and up to 454,450 controls) and separately for the subset of Europeans (about half as many strokes).^11^

### UK Biobank

The UK Biobank (UKB) is a prospective study of 502,506 men and women, aged 40–69 years living in the United Kingdom who were enrolled between 2006 and 2010.^9^ Details of the ethics approval, study methods, and baseline characteristics have been previously reported (Supplementary Methods 1 and 2).^9^ Participants were followed up for a mean of 8 years through linkages to death registries and hospital admission records. Criteria for diagnosis of ischaemic stroke cases (ICD-10: I63) were pre-specified and included both cases recorded prior to enrolment and incident cases recorded during follow-up (Supplementary Methods 3). Atrial fibrillation included admission to hospital (ICD-10: I48) and self-reported cases at baseline. Stroke cases with a history of atrial fibrillation prior to onset of stroke were classified as having presumed cardioembolic stroke. Genotyping using Affymetrix arrays with imputation into multiple reference panels (Supplementary Methods 4) was available for 483,420 participants passing quality control (Supplementary Methods 5). After exclusions for non-white British ancestry (n=78,674), and relatedness (n=67,201; kinship coefficient ≥0.125), a total of 336,750 UKB participants were included in the present genetic analyses (Supplementary Figure 1).

### China Kadoorie Biobank

The China Kadoorie Biobank (CKB) is a prospective study of 513,214 men and women, aged 30–79 years who were enrolled from 10 (5 urban and 5 rural) geographically defined regions of China between 2004 and 2008.^10^ Details of the ethics approval, study methods, and baseline characteristics have been previously reported (Supplementary Methods 6 and 7).^10^ Compared to participants in UKB, those in CKB were on average 5 years younger (mean age 51.6 [standard deviation 10.6] versus 56.4 [8.1] years), had lower mean levels of blood pressure, and were less highly educated (Supplementary Table 1). Participants were followed up for a mean of 10 years through linkages to death and stroke registries and health insurance claims records. Adjudication of stroke was undertaken by review of clinical findings from medical records and brain imaging reports (available for >92% of stroke cases with retrieved records) by specialist clinicians using a defined protocol (Supplementary Methods 8). Presumed cardioembolic strokes were identified from confirmed ischaemic stroke cases based on the Causative Classification System criteria.^13^ Other confirmed ischaemic stroke cases were further classified by brain infarct size into lacunar and other non-lacunar stroke subtypes. Data on atrial fibrillation were not systematically recorded at baseline or during follow-up in CKB, but electrocardiographic evidence of atrial fibrillation and other major and minor sources of cardioembolism were recorded by adjudicating physicians. Genotyping using Affymetrix arrays with imputation into the 1,000 Genomes reference panel (Supplementary Methods 9) was available for 100,706 participants passing quality control (Supplementary Methods 10), comprising a sample of 76,020 participants randomly selected from the CKB population and an additional 24,686 selected for nested case-control studies of incident cardiovascular or respiratory disease. After relatedness exclusions (n=28,233; kinship coefficient >0.05), the present genetic analyses involved 58,277 CKB participants (53,346 from the population-based subset, and 4,931 additional ischaemic stroke cases included only in analyses of ischaemic stroke outcomes) (Supplementary Figure 2).

### Height

Participants in CKB were on average shorter (10 cm in men, 8 cm in women: Supplementary Table 1) than those in UKB and the standard deviations (SDs) of directly-measured height in UKB and CKB, respectively, were 6.8 cm and 6.5 cm in men, and 6.3 cm and 6.0 cm in women. Separately in UKB and CKB, following methodology used in the Genetic Investigation of Anthropometric Traits (GIANT) Consortium, a measured height phenotype was constructed: within strata by sex (and by region in CKB) directly-measured height (Supplementary Methods 2 and 7) was adjusted for age and age^2^, and the residuals were transformed using an inverse normal transformation, yielding a measured height phenotype in study and sex-specific SD units. This transformed height phenotype (referred to as "height” or "measured height”) was used for all analyses (unless "directly-measured” is explicitly stated).

### Other anthropometric traits

Fat body mass was estimated as weight multiplied by percentage body fat measured by bio-impedance (Supplementary Methods 2 and 7). Lean body mass was estimated as weight minus fat body mass. Lung function measures (forced vital capacity [FVC] and forced expiratory volume in 1 second [FEV1]) were restricted to those with reliable values (Supplementary Methods 2 and 7; Supplementary Figure 2). Compared with UKB participants, those in CKB had lower mean levels of BMI (4 kg/m^2^ in men, 3 kg/m^2^ in women), and lean body mass (14 kg in men, 7 kg in women: Supplementary Table 1).

### Instruments for genetically-determined height

For MEGASTROKE, height-associated single nucleotide polymorphisms (SNPs) from the GIANT genome-wide association study (GWAS) report in 2018^14^ (which also included data from the whole of UKB) were used for both multiple and European ancestry analyses (Supplementary Table 2). Genetic instruments for a two-sample MR approach were constructed separately for UKB and CKB from findings in populations independent of the respective populations; for UKB, height-associated SNPs were obtained from an earlier (2014) GIANT study independent of UKB;^15^ for CKB, both the GIANT GWAS (2018)^14^ and a smaller GWAS from Biobank Japan,^16^ involving participants of East Asian ancestry, were used to optimise the genetic instrument for height.

The SNPs selected from these GWAS studies, together with their published single-variant effect sizes on height, were those associated with height at genome-wide significance available in the present studies (Supplementary Methods 11). The SNPs from each GWAS were linkage disequilibrium pruned (r^2^<0.2) using linkage disequilibrium estimates in UKB for GIANT and in CKB for Biobank Japan (i.e. where r^2^ between SNPs was ≥0.2, the SNP with the lowest p-value for association with height in the GWAS was retained). This left 2,931 and 678 SNPs for MEGASTROKE and UKB, respectively, and 2,812 and 536 SNPs from GIANT and Biobank Japan, respectively for CKB.

For UKB and CKB, genetic risk scores for each individual were constructed as the sum of the number of each height-associated effect alleles weighted by their published single-variant effect sizes on height (Supplementary Methods 11). For CKB, the genetic risk score was the unweighted average of genetic risk scores constructed from the GIANT (2018)^14^ and Biobank Japan^16^ height-associated SNPs (other weightings were assessed in sensitivity analyses but had less explanatory power [Supplementary Table 3]). The effects of SNPs on height in UKB and CKB estimated separately for each SNP using linear regression adjusted for age, age^2^, sex, region (in CKB only), genomic principal components (40 in UKB and 14 in CKB), and genotyping array type were also compared with the published effect sizes on height.

The genetic risk score in UKB explained 17.0% of the variance of height (Supplementary Methods 11; Supplementary Table 3) and the effect sizes of the SNPs in UKB were highly correlated with the effects in the source GWAS^15^ (r=0.94: Figure 1). In CKB, the genetic risk scores from GIANT, Biobank Japan and the average genetic risk score, respectively, explained 11.8%, 11.3% and 15.5% of the variance of height (Supplementary Table 3). SNP effect sizes in CKB were less strongly correlated with effects in GIANT (r=0.63),^14^ but were more strongly correlated with effects in Biobank Japan (r=0.90, respectively: Figure 1). One unit of the respective genetic risk score was associated with 0.84 SD of measured height in UKB and 0.99 SD in CKB.

**Figure 1:**
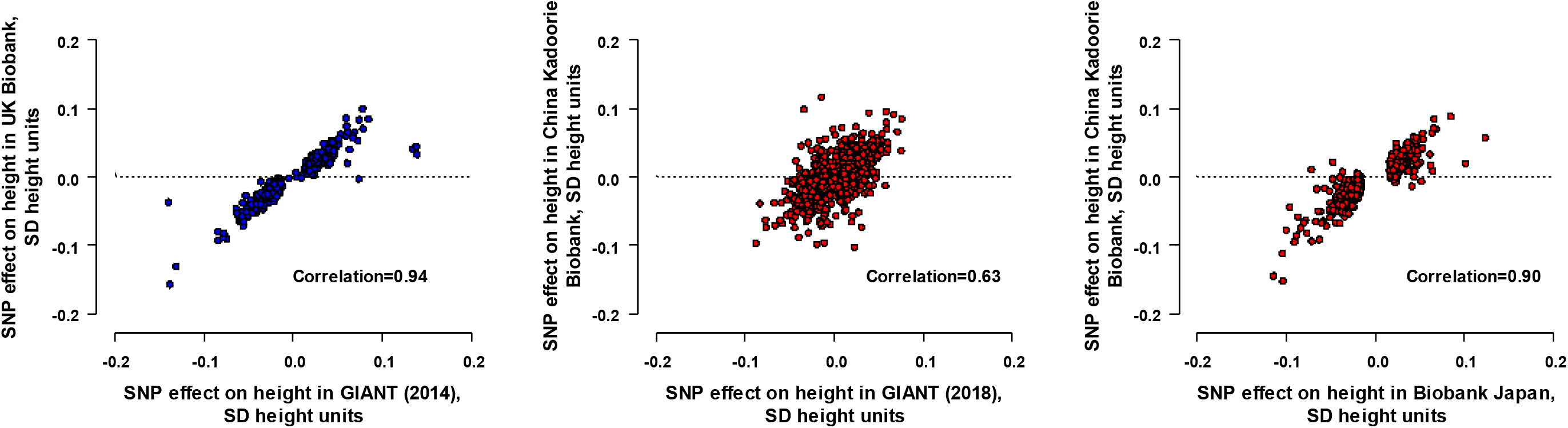
Effects of height-associated SNPs on height in UK Biobank and China Kadoorie Biobank. SD=standard deviation; SNP=single nucleotide polymorphism; GIANT=Genetic Investigation of Anthropometric Traits. For UK Biobank (336,750 participants), the effects on height were estimated for 678 SNPs from GIANT (2014)^15^. For China Kadoorie Biobank (53,346 participants), the effects on height were estimated for 2,812 SNPs from GIANT (2018)^14^ and 536 SNPs from Biobank Japan^16^. Each set of SNPs from GiAnT or Biobank Japan was linkage disequilibrium pruned (r^2^<0.2). The effect sizes on height were adjusted for age, age^2^, sex, region (in China Kadoorie Biobank only), genomic principal components, and genotyping array type. SNPs with minor allele frequencies of <0.005 were not shown. In UK Biobank, the genetic risk score explained 17.0% of the variance of height and, in China Kadoorie Biobank, the genetic risk scores from GIANT (2018)^14^, Biobank Japan^16^ and the average genetic risk score, respectively, explained 11.8%, 11.3% and 15.5% of the variance of height.

### Genetic analyses

For MEGASTROKE, causal effects were estimated by inverse-variance weighted random-effects SNP-level meta-analysis^17^ using GWAS summary results on stroke from MEGASTROKE^11^ (Supplementary Methods 12; Supplementary Figure 3). For UKB and CKB, the ratio method for single instruments was applied to the respective genetic risk scores to estimate the genetically-instrumented causal effects on outcomes per one standard deviation (1-SD) of measured height (Supplementary Methods 13). Specifically, logistic regression was used to assess associations of genetic risk scores with stroke outcomes (after adjustment for age, age^2^, sex, region in CKB, genomic principal components, and genotyping array type), and subsequently coefficients from these regressions were divided by the regression coefficient of measured height on the genetic risk score (0.84 SD of measured height in UKB and 0.99 SD in CKB).^17^ All effects presented as associations of genetically-determined height are the instrumented effects per 1-SD higher measured level of height (Supplementary Figure 3).

To investigate potential mediators of associations between height and ischaemic stroke, cross-sectional associations of genetically-determined height with established cardiovascular risk factors and anthropometric traits were assessed in UKB and CKB using linear or logistic regression as appropriate, with adjustment for age, sex, region in CKB, genomic principal components, and genotyping array type. For these cross-sectional associations, anthropometric traits and lung function were standardised (by dividing by their SD within each sex) in the UKB and CKB populations. The ratio method was then applied to regression results and, as for the disease outcomes, the genetically-instrumented effects presented. As, in large samples such as this study, t-statistics closely approximate z-statistics, the t-statistics for the associations of covariates in the linear regressions are referred to as z-statistics in this report. These were used to assess the strength and direction of the associations of height with cardiovascular and anthropometric factors to permit comparisons of z-statistics up to about ±500, which is beyond the convenient ranges for p-values (z-statistics of ±1.96 and of ±37 correspond to 2p=0.05 and 2p≈1×10^-300^, respectively).

### Sensitivity analyses

As MR inference relies on various assumptions, additional sensitivity analyses in MEGASTROKE included MR-Egger analyses to help assess any pleiotropic effects of height on other factors and MR-PRESSO analyses to correct for pleiotropy, if any, by removal of outliers (Supplementary Methods 14).^18^ As there is some overlap of the populations in MEGASTROKE with those in GIANT (2018)^14^ (Supplementary Methods 14) but not with UKB, the sensitivity analyses were repeated using effect sizes on height estimated in UKB.

### Observational analyses

Observational analyses were restricted to participants with no prior history of ischaemic heart disease or stroke in UKB (Supplementary Figure 1) and CKB (Supplementary Figure 2; Supplementary Methods 15). Hazard ratios (HRs) for the associations of measured height (grouped and as a linear term) with incident ischaemic stroke and ischaemic stroke subtypes post-recruitment, were estimated by Cox regressions stratified by age-at-risk (in 5-year groups), sex, and region (10 regions in CKB), with adjustment for possible baseline confounders (Supplementary Methods 15). Cross-sectional associations of measured height with cardiovascular and anthropometric factors at baseline were assessed using linear or logistic regression as appropriate, and adjusted for age (in 5-year groups), sex, year of birth, and region in CKB. All statistical analyses were conducted in SAS (version 9.4) and R (version 3.3.3).

## Results

Genetically-determined height was inversely associated with ischaemic stroke in MEGASTROKE in both multiple ancestries (odds ratio [OR]: 0.96; 95% CI: 0.93, 0.98) per 1-SD taller height, n=60,341 cases) and the European ancestry subset (Figure 2). The genetic associations in UKB and CKB were also consistent with the results in MEGASTROKE (Figure 2). However, the results for overall ischaemic stroke masked directionally opposing associations with different subtypes of ischaemic stroke.

**Figure 2:**
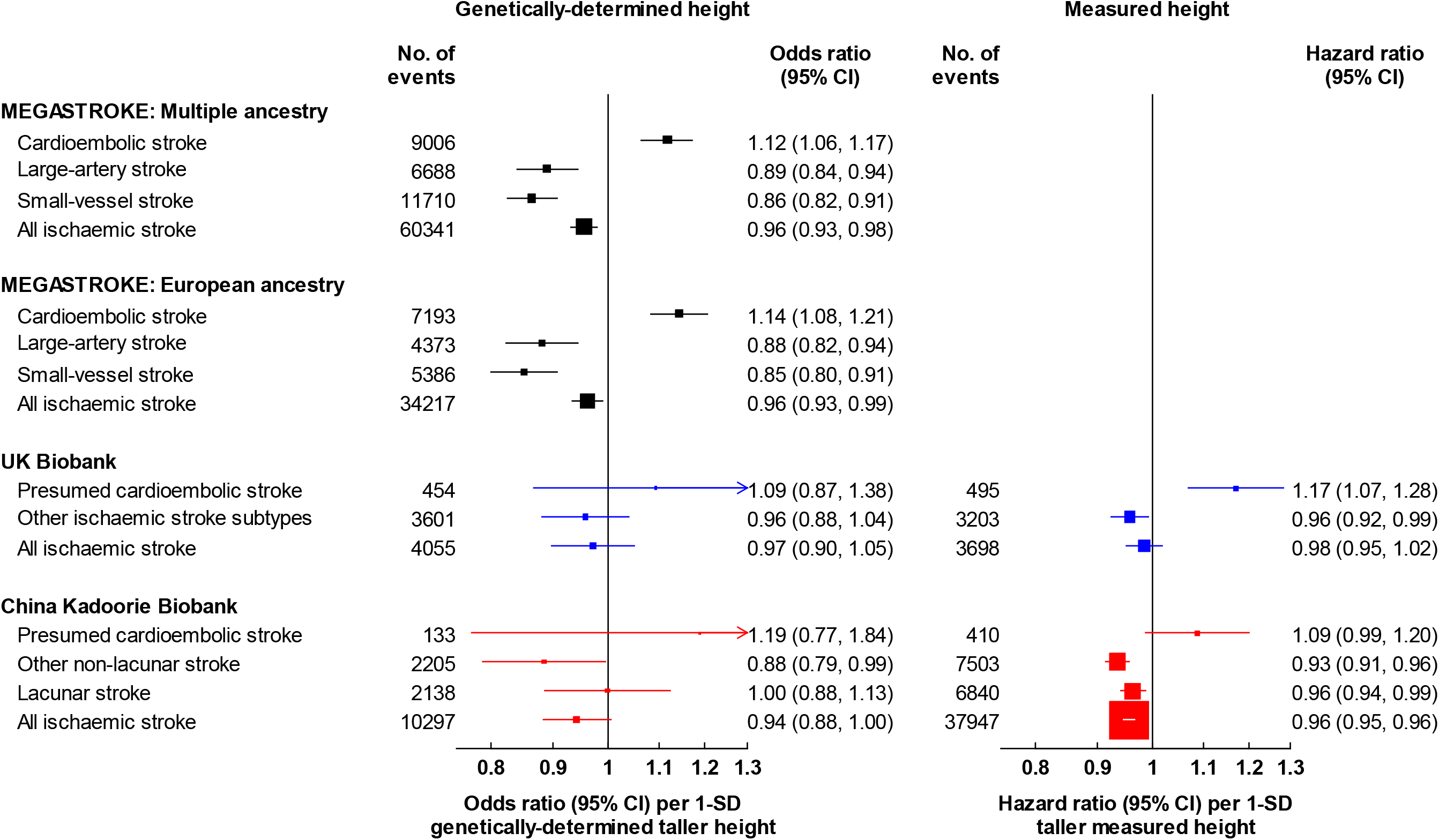
Associations of measured and genetically-determined height with ischaemic stroke and its subtypes in MEGASTROKE, UK Biobank, and China Kadoorie Biobank. SD=standard deviation. The numbers of events reported for MEGASTROKE were the maximum number of cases available in the genetic summary data. For UK Biobank and China Kadoorie Biobank, respectively, the SDs of directly-measured height were 6.8 cm versus 6.5 cm for men, and 6.3 cm versus 6.0 cm for women. Genetic associations in UK Biobank and China Kadoorie Biobank were adjusted for age, age^2^, sex, region (in China Kadoorie Biobank only), genomic principal components, and genotyping array type, and observational associations were stratified by age-at-risk (in 5-year groups), sex, and region (in China Kadoorie Biobank only), and adjusted for additional potential confounders (Supplementary Methods 15).

In MEGASTROKE, genetically-determined height was positively associated with cardioembolic stroke (OR per 1-SD taller height: 1.12 [95% CI: 1.06, 1.17], n=9,006), but was inversely associated with large-artery stroke (0.89 [0.84, 0.94], n=6,688) and small-vessel stroke (0.86 [0.82, 0.91], n=11,710) in multiple ancestries and were similar in the European ancestry subset (Figure 2). Similar findings were present in both UKB and CKB: for presumed cardioembolic stroke, the ORs were 1.09 (95% CI: 0.87, 1.38; n=454 cases) in UKB and 1.19 (0.77, 1.84; n=133 cases) in CKB; for other subtypes of ischaemic stroke, the corresponding ORs were 0.96 (95% CI: 0.88, 1.04; n=3,601) in UKB, while in CKB they were 0.88 (0.79, 0.99; n=2,205) for other non-lacunar stroke and 1.00 (0.88, 1.13; n=2,138) for lacunar stroke.

Sensitivity analyses in MEGASTROKE showed no evidence of directional pleiotropy for ischaemic stroke or its subtypes (p-values >0.15 for non-zero MR-Egger intercepts, Supplementary Table 4). The MR-PRESSO analyses identified only a few outlying SNPs (n≤3) and their exclusion had no impact on the causal estimates. There was no evidence of bias due to sample overlap as the causal estimates based on UKB effect sizes on height were largely unchanged.

Taller measured height was inversely and log-linearly associated with risk of ischaemic stroke in both UKB and CKB (Figure 3). The associations of measured height with ischaemic stroke subtypes in UKB and CKB were also consistent with the genetic associations in MEGASTROKE (Figure 2).

**Figure 3:**
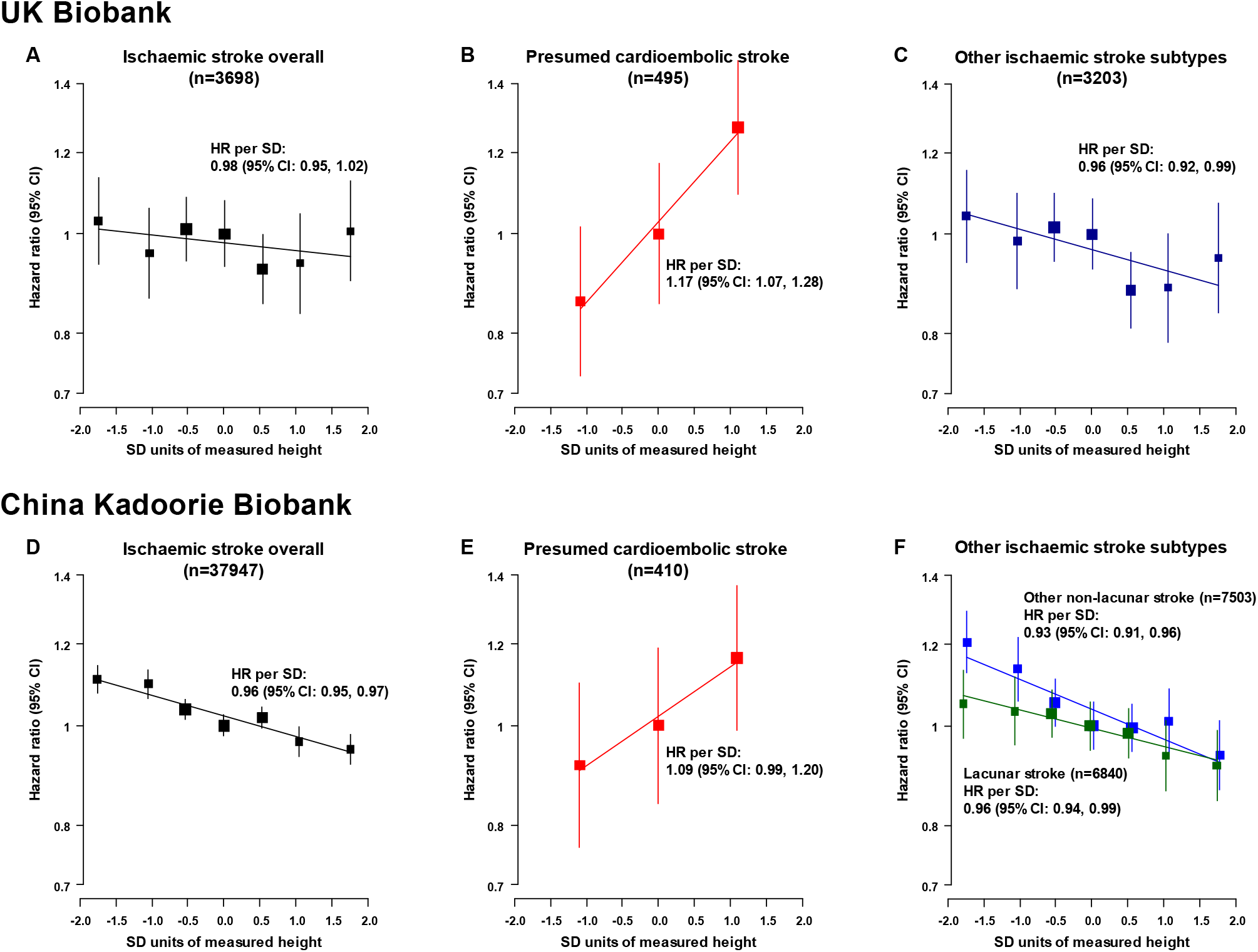
Associations of measured height with ischaemic stroke and its subtypes in UK Biobank and China Kadoorie Biobank. SD=standard deviation; HR=hazard ratio. For UK Biobank (482,928 participants) and China Kadoorie Biobank (490,067 participants), respectively, the SDs of directly-measured height were 6.8 cm versus 6.5 cm for men, and 6.3 cm versus 6.0 cm for women. HRs were stratified by age-at-risk (in 5-year groups), sex, and region (in China Kadoorie Biobank only), and adjusted for additional potential confounders (Supplementary Methods 15). Tenths of measured height were used to examine the shape of the associations of height with ischaemic stroke subtypes, except for presumed cardioembolic stroke where thirds were used due to the lower number of cases. When tenths of height were plotted, consecutive pairs of the middle 6 tenths were combined (to give 7 groups). HRs were presented as floating absolute risks relative to the middle height category (whereby standard errors were assigned approximately independently of each category to avoid restricting comparisons to any arbitrary reference groups).

The associations of genetically-determined and measured height with established cardiovascular risk factors and anthropometric traits are shown in Tables 1 and 2. The effects of genetically-determined height on risk factors were all directionally concordant and broadly consistent between UKB and CKB (Table 1; the generally lower z-statistics in the genetic comparisons in CKB reflect the smaller number of participants studied). Both genetically-determined and measured height were strongly associated with lean body mass (in UKB, 0.5–0.6 SD higher lean body mass per 1-SD taller genetically-determined height, z=98 in men, 87 in women) and with lung function (0.3–0.4 SD higher FEV1 or FVC, z=50–65).

**Table 1.** Associations of genetically-determined height with cardiovascular risk factors and anthropometric traits in UK Biobank and China Kadoorie Biobank.

| Baseline characteristic | UK Biobank (n=336750) |  | China Kadoorie Biobank (n=53346)* |  |
| --- | --- | --- | --- | --- |
|  | Effect (95% CI) per 1-SD genetically-determined taller height* | Z-statistic | Effect (95% CI) per 1-SD genetically-determined taller height* | Z-statistic |
| **Diagnosed prior disease (odds ratio)** |  |  |  |  |
| Diabetes | 0.96 (0.92, 1.00) | -2.0 | 0.99 (0.91, 1.09) | -0.2 |
| Atrial fibrillation | 1.33 (1.25, 1.42) | 8.8 | NA | NA |
| Hypertension | 0.91 (0.89, 0.93) | -9.9 | 0.97 (0.91, 1.04) | -0.7 |
| **Blood pressure (mmHg)** |  |  |  |  |
| Systolic blood pressure | -1.16 (-1.31, -1.01) | -15.6 | -0.10 (-0.53, 0.33) | -0.5 |
| Diastolic blood pressure | -0.44 (-0.53, -0.36) | -10.4 | -0.10 (-0.33, 0.14) | -0.8 |
| **Blood lipids** |  |  |  |  |
| LDL cholesterol (mmol/L) | -0.042 (-0.049, -0.034) | -11.0 | -0.054 (-0.102, -0.006) | -2.2 |
| HDL cholesterol (mmol/L) | -0.009 (-0.012, -0.005) | -5.4 | -0.008 (-0.030, 0.014) | -0.8 |
| Triglycerides (mmol/L) | -0.034 (-0.043, -0.026) | -7.8 | -0.042 (-0.153, 0.068) | -0.8 |
| Apolipoprotein B (g/L) | -0.014 (-0.016, -0.012) | -13.1 | -0.019 (-0.033, -0.004) | -2.6 |
| **Lung function (SD units within sex)** |  |  |  |  |
| FEV1, men | 0.316 (0.304, 0.329) | 50.3 | 0.222 (0.195, 0.249) | 16.4 |
| FEV1, women | 0.299 (0.288, 0.310) | 52.0 | 0.223 (0.202, 0.245) | 20.7 |
| FVC, men | 0.417 (0.404, 0.430) | 64.9 | 0.269 (0.241, 0.297) | 19.0 |
| FVC, women | 0.384 (0.372, 0.395) | 65.3 | 0.262 (0.240, 0.284) | 23.4 |
| **Anthropometric measures (SD units within sex)** |  |  |  |  |
| BMI, men | -0.071 (-0.084, -0.059) | -11.3 | -0.053 (-0.085, -0.020) | -3.2 |
| BMI, women | -0.072 (-0.083, -0.060) | -12.3 | -0.071 (-0.097, -0.046) | -5.6 |
| Waist-to-hip, men | -0.029 (-0.041, -0.017) | -4.7 | -0.036 (-0.070, -0.002) | -2.1 |
| Waist-to-hip, women | -0.026 (-0.037, -0.015) | -4.6 | -0.014 (-0.039, 0.011) | -1.1 |
| Weight, men | 0.396 (0.384, 0.408) | 63.7 | 0.338 (0.308, 0.368) | 22.4 |
| Weight, women | 0.323 (0.312, 0.335) | 55.8 | 0.335 (0.311, 0.358) | 27.8 |
| Lean body mass, men | 0.589 (0.577, 0.600) | 97.7 | 0.489 (0.459, 0.519) | 32.3 |
| Lean body mass, women | 0.493 (0.482, 0.504) | 86.9 | 0.594 (0.571, 0.617) | 51.3 |

**Table 2:** Associations of measured height with cardiovascular risk factors and anthropometric traits in UK Biobank and China Kadoorie Biobank.

| Baseline characteristic | UK Biobank (n=482928) |  | China Kadoorie Biobank (n=490067)* |  |
| --- | --- | --- | --- | --- |
|  | Effect (95% CI) per 1-SD taller measured height* | Z-statistic | Effect (95% CI) per 1-SD taller measured height* | Z-statistic |
| **Diagnosed prior disease (odds ratio)** |  |  |  |  |
| Diabetes | 0.85 (0.84, 0.86) | -28.5 | 1.06 (1.04, 1.07) | 8.4 |
| Atrial fibrillation | 1.31 (1.28, 1.34) | 22.2 | NA | NA |
| Hypertension | 0.87 (0.87, 0.88) | -41.6 | 1.13 (1.12, 1.14) | 24.8 |
| **Blood pressure (mmHg)** |  |  |  |  |
| Systolic blood pressure | -1.16 (-1.21, -1.11) | -46.4 | 0.32 (0.27, 0.38) | 11.7 |
| Diastolic blood pressure | -0.36 (-0.39, -0.33) | -25.3 | 0.44 (0.41, 0.47) | 28.5 |
| **Blood lipids** |  |  |  |  |
| LDL cholesterol (mmol/L) | -0.017 (-0.019, -0.014) | -13.0 | 0.008 (-0.001, 0.018) | 1.7 |
| HDL cholesterol (mmol/L) | 0.005 (0.004, 0.007) | 10.2 | -0.012 (-0.016, -0.008) | -5.8 |
| Triglycerides (mmol/L) | -0.047 (-0.050, -0.044) | -31.7 | 0.049 (0.026, 0.071) | 4.3 |
| Apolipoprotein B (g/L) | -0.009 (-0.009, -0.008) | -24.6 | 0.003 (0.000, 0.006) | 2.3 |
| **Lung function (SD units within sex)** |  |  |  |  |
| FEV1, men | 0.370 (0.366, 0.374) | 173.7 | 0.281 (0.278, 0.284) | 168.7 |
| FEV1, women | 0.355 (0.352, 0.359) | 188.1 | 0.280 (0.277, 0.282) | 197.9 |
| FVC, men | 0.452 (0.448, 0.456) | 216.4 | 0.311 (0.308, 0.315) | 186.6 |
| FVC, women | 0.425 (0.421, 0.429) | 227.3 | 0.305 (0.302, 0.307) | 215.1 |
| **Anthropometric measures (SD units within sex)** |  |  |  |  |
| BMI, men | -0.056 (-0.060, -0.051) | -25.8 | 0.040 (0.036, 0.044) | 19.1 |
| BMI, women | -0.119 (-0.123, -0.115) | -61.9 | 0.002 (-0.002, 0.005) | 0.9 |
| Waist-to-hip, men | -0.048 (-0.052, -0.044) | -22.6 | 0.043 (0.039, 0.047) | 19.9 |
| Waist-to-hip, women | -0.075 (-0.079, -0.071) | -39.3 | 0.003 (-0.001, 0.006) | 1.5 |
| Weight, men | 0.410 (0.406, 0.414) | 208.9 | 0.436 (0.433, 0.440) | 253.2 |
| Weight, women | 0.274 (0.270, 0.277) | 146.9 | 0.407 (0.404, 0.410) | 263.7 |
| Lean body mass, men | 0.608 (0.605, 0.611) | 363.3 | 0.586 (0.583, 0.589) | 384.1 |
| Lean body mass, women | 0.470 (0.466, 0.473) | 276.8 | 0.650 (0.648, 0.652) | 555.4 |

Genetically-determined taller height was also associated with lower levels of LDL cholesterol, HDL cholesterol, and blood pressure in both UKB and CKB, but the estimated effect sizes on blood pressure were greater in UKB than in CKB (−1.16 versus −0.10 mmHg). In UKB, the findings for measured and genetically-determined height with systolic blood pressure were highly consistent (Tables 1 and 2), but in CKB the measured height was positively, rather than inversely, associated with systolic blood pressure, suggesting that this association might reflect confounding in CKB. Both genetically-determined and measured height were strongly positively associated with atrial fibrillation at baseline (available only in UKB) with ORs per 1-SD taller height of 1.33 (95% CI: 1.25, 1.42) and 1.31 (95% CI: 1.28, 1.34), respectively (Tables 1 and 2).

## Discussion

In this large MR study of ischaemic stroke involving multiple ancestries, there were modest inverse associations of both genetically-determined and measured height with overall ischaemic stroke. However, these masked much stronger directionally opposing associations of height with cardioembolic versus other ischaemic stroke subtypes. In MEGASTROKE (multiple ancestries), a 1-SD genetically-determined taller height was associated with 12% higher risk of cardioembolic stroke, but with 11% and 14% lower risks of large-artery stroke and small-vessel stroke, respectively. The associations of both genetically-determined and measured height with ischaemic stroke subtypes in UKB and CKB were concordant with each other and with the genetic associations in MEGASTROKE. The highly consistent findings from observational and MR approaches across three different studies provide support for the causal relevance of the associations of height with ischaemic stroke subtypes.

The association of 1-SD taller genetically-determined height was associated with a 11% lower risk of large artery stroke in the present study and consistent with previously reported associations with ischaemic heart disease (OR per 1-SD taller genetically-determined height: 0.88 [95% CI: 0.82, 0.95]).^3^ These associations of genetically-determined height may be mediated in part by established causal risk factors for cardiovascular disease.

LDL cholesterol has been shown to be causally associated with increased risk of ischaemic stroke in both European and Chinese populations,^19^ with the strongest association observed with large-artery stroke and little association seen with cardioembolic stroke.^20^ Thus, the inverse association of genetically-determined height with LDL cholesterol levels in both UKB and CKB could explain some of the inverse associations of height with large-artery stroke and to a lesser extent, with small-vessel stroke, although the mechanism by which height might do this is unclear. Genetically-determined taller height was also associated with lower mean levels of blood pressure in both studies (about 1 mmHg lower in UKB, but only 0.1 mmHg in CKB: Table 1); based on the UKB effect this would be expected to translate to about 3% lower risk of ischaemic stroke and 2–5% lower risk of each ischaemic stroke subtype.^21^ By contrast with the consistency of the genetic associations, the observational associations were not as consistent between UKB and CKB, possibly reflecting residual confounding (e.g., by socioeconomic factors, as blood pressure and height are positively correlated with income in China^22^) or reverse causality (e.g. from use of LDL-lowering drugs), illustrating the advantage of MR analyses.

The associations of height with ischaemic stroke subtypes may reflect a direct causal effect of body dimensions on stroke subtypes, or the effects of some other correlated anthropometric trait (such as lean body mass) on the diseases. Previous MR studies have suggested that greater lung function may act as a possible mediator of the protective effect of height on ischaemic heart disease.^4^ In both UKB and CKB, taller height was associated with higher lung function and so lung function could account for some of the protective effects of height but is unlikely to explain the adverse associations of height with cardioembolic stroke.^4^

Greater lean body mass is the chief anthropometric risk factor (stronger than height) for atrial fibrillation^6^ and previous MR studies have confirmed its causal relevance for atrial fibrillation.^5^ Atrial fibrillation is a major risk factor for cardioembolic stroke,^23^ and the present study provides support for the causal relevance of taller height with the more severe outcome of cardioembolic stroke. Larger left atrial diameter, present in taller people, has also been associated with higher risks of atrial fibrillation and embolism from cardiac sources,^24^ but whether these associations are mediated by lean body mass or some other aspect of body dimensions has not been previously studied. Higher levels of lean body mass have also been positively associated with carotid intima-media thickness, left ventricular mass and cardiac wall thickness, but not with atherosclerosis.^25^

Genotypes associated with height, education, blood pressure and several chronic diseases have been shown to be correlated within spouse-pairs (i.e., indicate assortative mating) which can lead to indirect effects of genotypes in offspring, in violation of MR assumptions.^26^ Family-based studies have reported that such indirect genetic effects of non-transmitted alleles could explain about 12% of the genetic effect on height.^27^ As desirable traits such as higher income, taller height and healthy traits tend to cluster in mates, assortative mating could explain some of the protective associations of taller height, but is unlikely to explain the adverse associations of height with atrial fibrillation and cardioembolic stroke.

The opposing associations of height with cardioembolic and other ischaemic stroke subtypes highlight the importance of considering ischaemic stroke subtypes as distinct diseases. Many studies (e.g. UKB) do not currently have detailed and reliable ischaemic stroke subtyping, limiting their use for causal inference. Studies examining the associations of risk factors with overall ischaemic stroke may incorrectly estimate medically relevant associations of some risk factors with individual ischaemic stroke subtypes. Subtyping is also important in clinical practice for prevention of stroke recurrence, where the impact of treatments, such as statins or anticoagulants may vary in different ischaemic stroke subtypes.^20^

Men and women in CKB were 10 and 8 cm shorter (about 1.5 SD), respectively, than their counterparts in UKB (Supplementary Table 1). If the MR associations in Figure 2 are causal, this would translate to adults in China having a higher risk of some ischaemic stroke subtypes (particularly for large-artery stroke and small-vessel stroke subtypes), a lower risk of cardioembolic stroke, and a modest lower risk of all ischaemic stroke through shorter stature compared with Europeans. Hence, initiatives to improve childhood nutrition, resulting in an increase in stature, could reduce the net risk of ischaemic stroke in adult life. In Japan, mean adult height has increased by about 8 cm between the 1960s and 2000s, and so may have contributed to some of the 70% decline in stroke incidence observed during this period,^28^ which is not fully explained by trends in blood pressure control.^29^

This was the first large genetic study to examine the associations of height with ischaemic stroke subtypes and risk factors in multiple ancestries. Genetic associations in different ancestry groups were supported by observational associations, and different methods were used to check MR assumptions. Nevertheless, the present study also had several limitations: firstly, there were differences in the methodology for classification of ischaemic stroke subtypes and a lack of reliable subtyping in all of the populations studied. As cardioembolic stroke has been reported to account for more than half of ischaemic stroke cases in some Western populations,^30^ the relatively low number of presumed cardioembolic stroke cases observed in both UKB and CKB is likely to be an underestimate of the true incidence of cardioembolic stroke subtypes. Secondly, although MR analyses avoid many of the biases (confounding and reverse causation) inherent in observational studies and sensitivity analyses exploring the robustness of the analyses to some of the assumptions that MR analyses rely on demonstrated no appreciable differences, violations due to assortative mating cannot be excluded. Thirdly, height has been estimated to have a SNP-based heritability of about 50% in both Europeans^15^ and East Asians^16^. The genetic risk scores for height used in UKB (based on an independent largely European ancestry-based GWAS) explained 19.7% of the variance in height in UKB, but the genetic risk score used in CKB (based on a large GWAS of height in a European population^14^ and a smaller GWAS of height in a Japanese population)^16^ explained only 15.5% of the variance in height in CKB. The present multiple ancestry analysis in MEGASTROKE may therefore have under-estimated the causal effects of height if the (European ancestry-derived) genetic risk score used was associated with smaller differences in height in the non-European ancestry populations.

Overall, the present genetic studies provide novel and reliable findings about the causal relevance of taller adult height for higher risks of atrial fibrillation and cardioembolic stroke, and lower risks of other ischaemic stroke subtypes. These findings provide further support for considering ischaemic stroke subtypes as distinct diseases in both clinical practice and research.

## Author contributions

All authors were involved in study design, conduct of the study, long-term follow-up, analysis of data, interpretation of results, or writing the report. SP had access to all the data in the study and had final responsibility for the decision to submit for publication.

## Data Availability

MEGASTROKE data are publicly available. UK Biobank data are available on application. China Kadoorie Biobank data sharing will be considered in line with the Nuffield Department of Population Health, University of Oxford, Data Access and Sharing Policy available at https://www.ndph.ox.ac.uk/files/about/ndph-data-access-policy-1.pdf

https://www.megastroke.org/

## Acknowledgements

The China Kadoorie Biobank study is jointly coordinated by the University of Oxford and the Chinese Academy of Medical Sciences. The baseline survey was supported by the Kadoorie Charitable Foundation, Hong Kong, China and the long-term continuation of the study was supported by the UK Wellcome Trust (202922/Z/16/Z, 104085/Z/14/Z, 088158/Z/09/Z), Chinese National Natural Science Foundation (81390540, 81390541, 81390544), and the National Key Research and Development Program of China (2016YFC0900500, 2016YFC0900501, 2016YFC0900504, 2016YFC1303904). The Clinical Trial Service Unit, University of Oxford, also acknowledges support from the UK Medical Research Council (MC_UU_00017/1, MC_UU_00017/5), the British Heart Foundation (CH/1996001/9454), the British Heart Foundation Oxford Centre for Research Excellence (RE/18/3/34214), and Cancer Research UK (C500/A16896). ABL was supported by the Clarendon Fund and by a Nuffield Department of Population Health Early Career Research Fellowship. JCH was supported by a British Heart Foundation Grant (FS/14/55/30806). The MEGASTROKE project received funding from sources specified at https://www.megastroke.org/acknowledgements.html. The research included in the present report used data obtained from the UK Biobank resource under application 10,061.

